# Socioeconomic status and severe mental disorders: A Bidirectional Multivariable Mendelian Randomisation Study

**DOI:** 10.1101/2023.05.25.23290516

**Authors:** Álvaro Andreu-Bernabeu, Javier González-Peñas, Celso Arango, Covadonga M. Díaz-Caneja

## Abstract

**Importance:** Despite the evidence supporting the relationship between socioeconomic status (SES) and severe mental disorders (SMD), the directionality of the associations between income or education and mental disorders is still poorly understood.

**Objective:** To investigate the potential bidirectional causal relationships between genetic liability to the two main components of SES (i.e., income and educational attainment (EA)) on three SMD: schizophrenia (SCZ), bipolar disorder (BD) and major depressive disorder (MDD) using multivariable Mendelian randomisation.

**Design, Setting, and Participants:** We performed a bidirectional, two-sample multivariable Mendelian randomisation (MVMR) study using summary statistics from the latest genome-wide associations studies (GWAS) of the Psychiatric Genomics Consortium (PGC), the UK biobank and the Social Science Genetic Association Consortium (SSGAC) to dissect the potential associations of income and EA with schizophrenia, BD and MDD including data for participants of European ancestry only. Data covered September 2022 to January 2023.

**Main Outcomes and Measures:** Socioeconomic status phenotypes (household income from the UK biobank, n=397,751, and EA based on the largest meta-analysis GWAS of individuals of European ancestry, n=766,345) and SMD from the PGC (schizophrenia, n=127,906; BD, n=51,710; and MDD n=500,119). Follow-up analyses were performed using the intelligence meta-analytical phenotype, n=269,867.

**Results:** GWAS data were derived from participants of European ancestry included in the UK Biobank, SSGAC and PGC consortiums (samples ranging from 766,345 to 51,710 individuals). Univariable MR (UVMR) showed that genetic liability to household income was associated with decreased risk of schizophrenia (IVW-ORSCZ=0.58 per-one-SD increase; P-value= 0.016) and MDD (IVW-ORMDD:0.66 P-value: 9.81e-08), with a smaller reverse effect of schizophrenia and MDD on income (IVW-βSCZ= -0.1147, P-value=7.55E-08; IVW-βDEP=-0.0356, P-value=8.70E-04). Direct effects (MVMR) after adjusting for EA were comparable. UVMR showed that genetic liability to EA was associated with lower risk of MDD and higher risk of BD, with no significant effects on schizophrenia. After accounting for income in the MMVR analyses, direct effects of genetic liability to EA was associated with increased risk of BD (MVMR-ORBIP: 2.69 per SD increase, P-value= 0.0000113) and schizophrenia (MVMR-ORSCZ: 2.09 per-SD-increase, P-value= 0.00108), but not with MDD. Effects for the association of EA with schizophrenia and BD were larger when including genetic liability to intelligence in the MVMR model, thus suggesting that they might reflect a non-cognitive component.

**Conclusions and Relevance:** The findings of this multivariable Mendelian randomisation analysis suggest an heterogenous pattern of causal links between SES and SMD. We found evidence for a negative bidirectional association between genetic liability to household income and the risk of schizophrenia and MDD. On the contrary, we found a positive bidirectional relationship of genetic liability to EA with schizophrenia and bipolar disorder, which only becomes apparent after adjusting for income. These findings shed light on the directional mechanisms between social determinants and mental disorders and may help to guide public mental health strategies addressing inequality and economic disadvantages.

## Introduction

Social determinants and their potential aetiological role in mental disorders have been a major topic since the inception of psychiatry.^1^ Socioeconomic status (SES) plays a fundamental role in the performance of an individual in society, affecting well-being, longevity and health.^2,3^ Extensive evidence based on survey data, large cohort observational studies and family-level studies supports the association between mental disorders and lower income, disadvantaged socioeconomic position and homelessness.^4–6^ Recent work has also reported an association of parental socioeconomic status with offspring mental disorders such as major depressive disorder(MDD)^7^ or schizophrenia.^8^ However, the specific pattern of relationships is heterogeneous, as some disorders such as ASD or bipolar disorder (BD) are associated with higher parental or individual socioeconomic status,^9,10^ while depression or schizophrenia are associated with lower levels of education and income.^8,11–13^

Two classic theories have attempted to explain the nature of the relationship between SES and mental health: health selection and social causation. The health selection theory posits that individuals with mental disorders might be predisposed to reduced educational attainment (EA) and lower income. The social causation theory, in contrast, posits that social inequality, health resources gaps and context-related stress may increase the risk of mental disorders.^6,14–16^ The general consensus to date suggests that health selection could play a stronger role in severe mental disorders (SMD) like schizophrenia, whereas social causation would be more relevant for common mental disorders such as anxiety or depressive symptoms.^17,18^

Although researchers have considered socio-economic factors mainly environmental, the two core phenotypes comprising SES, income and educational attainment (EA), are substantially heritable (40-60%).^19,20^ Likewise, family and twin studies have estimated that genetics account for a large portion of the phenotypic variation of mental disorders, with heritability estimates of 65-80% in schizophrenia^21^ and 30-40% for MDD.^22^

Genome-wide association studies (GWAS) have supported observational evidence of heterogeneous patterns of genetic correlations between mental disorders and income.^23–25^ For example, two recent studies using Conditional Genome-wide Complex Trait Analysis (GCTA) and Genomic Structural Equation Modelling (GSEM) found how genetic associations among mental disorders were influenced by genetic overlap with SES traits,^26,27^ highlighting the polygenic and pleiotropic links between mental disorders and SES.

Despite the efforts to disentangle the complex relationship between SES and mental disorders, there is still ongoing debate due to the disparity of results and lack of replication.^8,17,28^ Further, conventional observational studies are affected by unmeasured confounders that preclude adequate assessment of the directional associations between SES and mental disorders.^11^ The use of univariable Mendelian randomisation (UVMR) has become popular for analysing causality between exposures to risk or protective factors and outcomes such as diseases in recent years, to overcome the limitations of observational studies.^29,30^ An extension of MR called multivariable Mendelian randomisation (MVMR) has recently been developed to allow the analysis of the independent causal effects of correlated exposures simultaneously.^31^

We performed bidirectional two-sample MR to determine whether genetic liability to household income and educational attainment were causally linked to three SMD (i.e., schizophrenia (SCZ), bipolar disorder (BD) and major depressive disorder (MDD)) with onset in adolescence or young adulthood and differing effects across the psychotic, cognitive and affective spectra. We used MVMR to disentangle the independent contribution of income and EA to SMD and the independent reverse effects of SMD on each SES trait.

## Methods

### Study design

Mendelian randomisation (MR) is an instrumental variable analysis method based on genetic variants (single nucleotide polymorphisms, SNPs). MR enables causal estimates between exposure and an outcome thanks to the natural randomisation of genetic variants that takes place in meiosis.^29^ MVMR is an extension of the MR method that allows the inclusion of instrumental variables of different exposures that are phenotypically and genetically associated in the same model in order to obtain the independent direct effects of each exposure on the outcome.^32^ We performed bidirectional two-sample UVMR and MVMR analyses to assess the causal links between SES (income and EA) and SMD in public datasets of European population. We performed follow-up analyses including genetic liability to intelligence^33^ in MVMR models to assess the causal links independently of intelligence. For more information on MR **see eMethods** and MR guidelines.^29,30,34^

## Data sources

### Socioeconomic Status Phenotypes

We used the largest publicly available genome-wide association study (GWAS) summary statistics of household income from the UK Biobank(n= 397,751)^35^. For EA we used the largest available GWAS from European Ancestry (n=766,345)^24^. The EA phenotype was a meta-analysis of 70 GWAS with standardised measures of years of schooling, including 442,183 individuals from the UK Biobank cohort.

### Mental Disorders

We selected the latest GWAS from the Psychiatric Genomics Consortium (PGC) for (i) SCZ (Wave-3, only European ancestry,^36^ comprising 52,017 cases and 75,889 controls); (ii) BD^37^, 20,352 cases and 31,358 controls; and (iii) MDD^38^, 170,756 cases and 329,443 controls.

### Intelligence

We used the largest GWAS meta-analysis of intelligence comprising 14 independent epidemiological cohorts of European ancestry (n= 269,867). The ethics declarations for each public dataset used in the present research can be found in their original publications.

## Genetic instruments and statistical analysis

We extracted genome-wide significant SNPs as instrumental variables at p < 5 × 10−8. For each exposure, we calculated the F statistic to analyse the instrument’s strength^39^ and used the rule of thumb of F>10 adopted in previous studies to define weak instrument bias.^30,34^

### Univariable Mendelian Randomisation

We conducted a bidirectional two-sample UVMR analysis to assess putative causality between household income and EA and the three SMD. We used an inverse-variance weighted method (IVW) as the main analysis, as recommended by available guidelines due to its robustness.^30^ However, as IVW may generate biased results in case of heterogeneity or violation of any assumption, we used Weighted median (WM),^40^ MR-Egger,^41^ Simple mode (SM),^42^ Weighted mode (WMo),^42^ heterogeneity tests and horizontal pleiotropy tests as sensitivity analyses.^41^ Additionally, we conducted Mendelian Randomisation Pleiotropy RESidual Sum and Outlier (MR-PRESSO) analysis which enables correction of horizontal pleiotropy via outlier removal.^43^ All analyses were performed using the “TwoSampleMR” package v 0.5.6 ^39^, with R version 4.1.2.

### Multivariable Mendelian Randomisation

The total effects provided by a classical UVMR method can suffer from bias when there is a high correlation between exposures, as is the case of income and EA. Hence, we evaluated i) the direct effect independent of EA of genetic liability to income on each SMD, and the direct effect of genetic liability to each SMD on income and ii) the direct effect independent of income of genetic liability to EA on each SMD, and the direct effect of genetic liability to each SMD on EA. Finally, we conducted post-hoc analyses including genetic liability to intelligence in the MVMR models to assess the association of EA and income with SMD independently of intelligence.

For each MVMR analysis, we applied the same LD-Clumping, harmonized method and palindromic SNPs exclusion as for UVMR using the “TwosampleMR” package.^39^ We then followed Sanderson et al (2021)^32^ to estimate causal effects and to correct for potentially weak and pleiotropic instruments through various sensitivity analyses. We estimated the instrumental variable strength using conditional-F-statistics and heterogeneity as well as horizontal pleiotropy using a modified Cochran’s Q-statistic. We performed IVM-MVMR regression of each model and robust-to-weak-instrument Q-statistic minimization, using non-parametric 1000 iteration bootstrap permutations. All MVMR analyses were performed with the MVMR package (more details in eMethod and [https://github.com/WSpiller/MVMR]).

### Interpretation of the estimates

For the analyses exploring the effect of genetic liability to income and EA on the risk of SMD, causal effects and 95% confidence intervals (CI) we expressed the increase in the odds of developing each mental disorder in standard deviations (SD). Analyses exploring the effect of SMD on EA and income were first multiplied by the SD of income(SD= 33,181£)^44^ and EA(SD=4.2 years of education)^24^ to convert them to British pounds and months and then expressed as the effect of a doubling (2-fold increase) of genetic liability to each SMD on the odds of income and EA, as recommended in previous studies.^45–47^ Finally, we defined significance at a false discovery rate (FDR) of 0.05 for each UVMR analysis. For the MVMR analyses we considered significant results included in the CI after non-parametric 1000-iteration bootstrap permutation. For more details see eMethods.

## RESULTS

### Effect of genetic liability to socioeconomic status on the risk of SMD

The results of the bidirectional UVMR (total effects) and MVMR (direct effects) analysis of genetic liability to income and EA on SMD are shown in Figures 1A and 1B.

#### Effects of genetic liability to educational attainment on mental disorders

Using the inverse-variance weighted (IVW) method, genetic liability to EA was univariately associated with a lower risk of MDD per SD-increase-unit (∼4.2 years schooling) (IVW-ORDEP=0.78, P-value=1.89E-10) and a higher risk of BD (IVW-ORBIP= 1.84, P-value=4.81E-09), but not with schizophrenia risk (IVW-ORSCZ=1.11, P-value= 0.263) Sensitivity analyses mirrored the results of the IVW method and egger-intercept showed non-significant results indicating that there was little evidence for directional genetic pleiotropy(eTable 2A,2B). After accounting for income, genetic liability to EA was associated with an almost 2.7-fold increased risk of BD (MVMR-ORBIP=2.69, P-value=0.0000113), and a 2-fold increased risk of schizophrenia (MVMR-ORSCZ=2.09, P-value=0.00108), with no significant effects on MDD risk. These results were consistent with the corresponding confidence intervals of the robust-to-weak instruments MVMR sensitivity analyses (see Figure 1A and eTable1 and 2 for more details)

**Figure 1:**
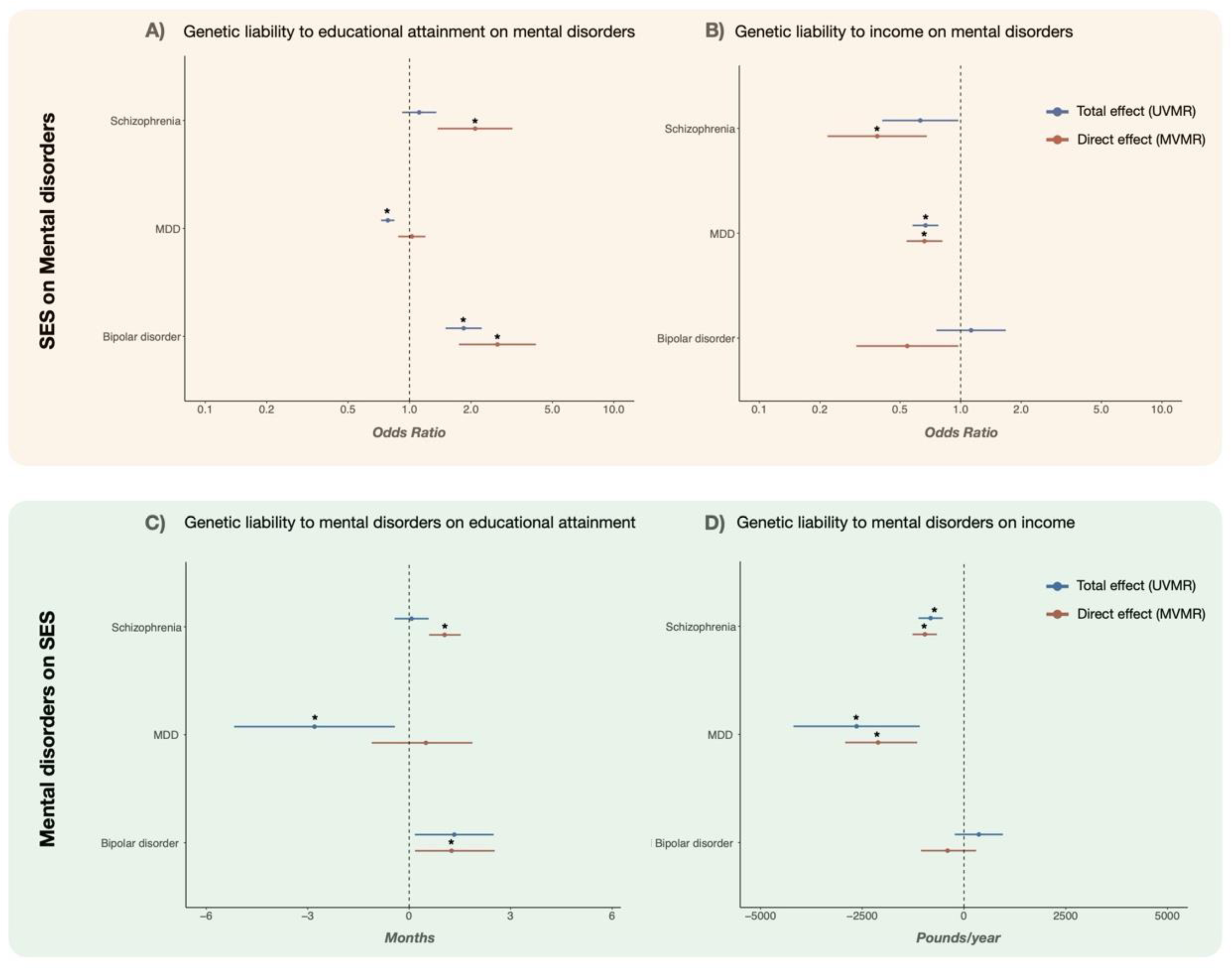
Directional associations between socioeconomic status (SES) and mental disorders: A: Total (UVMR, blue) and direct effects (MVMR, red) of genetic liability to educational attainment on the risk of severe mental disorders (SMD). B: Total (UVMR, blue) and direct effects (MVMR, red) of genetic liability to household income on the risk of SMD. C: Total (UVMR, blue) and direct effects (MVMR, red) of genetic liability to SMD on EA D: Total (UVMR, blue) and direct effects (MVMR, red) of genetic liability to SMD on household income. MR estimates and 95% confidence intervals (CI) for the effects of genetic liability to EA and income on the risk of SMD (1A,1B) per one-SD increase in each SES component (i.e., household income 33,181 pounds, EA 4.2 education years) on the odds of each SMD. For analysis investigating the effects of genetic liability of mental disorders to SES (1C,1D), estimates are provided per doubling (2-fold increase) of the prevalence of the exposure. Significant effects after false discovery rate (FDR) correction (pFDR <lt; 0.05) for total effects and after non-parametric 1,000 iteration bootstrap permutations for direct effects are marked with an asterisk. UVMR: Univariable Mendelian Randomisation, MVMR: Multivariable Mendelian Randomisation, EA: Educational Attainment, SES: Socioeconomic status. MDD: Major Depressive Disorder

#### Effects of genetic liability to household income on mental disorders

Genetic liability to household income was associated with lower risk of schizophrenia (IVW-ORSCZ=0.58 per-SD-increase, P-value=0.016) and MDD (IVW-ORMDD=0.66 per-SD-increase, P-value=9.81e-08). We found heterogeneity for both disorders, but there was limited evidence of horizontal pleiotropy, as suggested by the MR-Egger intercept (see eTable1 and 3). Using MVMR, the estimated direct effect of genetic liability to income, independent of EA, was nearly comparable to the total effect for both disorders (MVMR-ORSCZ: 0.32 per-SD-increase, P-value=0.00042; MVMR-ORMDD: 0.66, P-value=9.96E-05). We found no significant associations between income and BD in the UVMR or MVMR analyses (see Figure 1B and eTable1 and 3).

### Effect of genetic liability to SMD on socioeconomic status

The results of the bidirectional UVMR and MVMR analysis of the effects of genetic liability to schizophrenia, MDD and BD on income and EA are shown in Table 1 and Figures 1B and 1C.

**Table 1.A.** Estimates of the total and direct effects of genetic liability to mental disorders on genetic liability to income

| exposure | UVMR (IVW) \| Total Effect \|<br>Outcome: Income |  |  |  |  | MVMR \| Direct Effect (mental disorder not mediated by EA) \|<br>Outcome: Income |  |  |  |  |
| --- | --- | --- | --- | --- | --- | --- | --- | --- | --- | --- |
|  | nsnp | beta | SE | 95% CI | pval | nsnp | beta | SE | 95% CI* | pval |
| Schizophrenia | **149** | **-0.0356** | **0.0089** | **-0.048, -0.022** | **7.55E-08** | **329** | **-0.0417** | **0.006** | **-0.055, -0.029** | **5.27E-09** |
| Bipolar disorder | 13 | 0.0159 | 0.0131 | -0.0098, 0.041 | 0.2254 | 298 | -0.0175 | -0.0167 | -0.046, 0.013 | 0.09 |
| MDD | **44** | **-0.1147** | **0.0344** | **-0.182, -0.047** | **8.70E-04** | **304** | **-0.0918** | **-0.0849** | **-0.127, -0.05** | **8.61E-05** |

**Table 1.B.** Estimates of the total and direct effects of genetic liability to mental disorders on genetic liability to educational attainment

| exposure | UVMR (IVW) \| Total Effect \|<br>Outcome: Educational Attainment |  |  |  |  | MVMR \| Direct Effect (mental disorder not mediated by income) \|<br>Outcome: Educational Attainment |  |  |  |  |
| --- | --- | --- | --- | --- | --- | --- | --- | --- | --- | --- |
|  | nsnp | beta | SE | 95% CI | pval | nsnp | beta | SE | 95% CI* | pval |
| Schizophrenia | 145 | 0.002 | 0.007 | -0.012, 0.016 | 0.7785 | **164** | **0.0302** | **0.007** | **0.017, 0.044** | **5.78E-06** |
| Bipolar disorder | **13** | **0.038** | **0.017** | **0.004, 0.072** | **0.0247** | **55** | **0.0362** | **0.014** | **0.005, 0.073** | **8.17E-03** |
| MDD | **44** | **-0.0807** | **0.035** | **-0.149, -0.012** | **0.0211** | 77 | 0.0143 | 0.025 | -0.032, 0.054 | 0.63 |

*Abbreviations: nsnp: Number of single nucleotid polymorphism, MDD: Major Depressive disorder, CI: Confidence Interval, 95% CI\*: 95% confidence intervals included after non-parametric 1,000 iteration bootstrap permutations. Bold (pFDR < 0.05 and MVMR with 95% confidence intervals included after non-parametric 1000 permutations)*

#### Effects of genetic liability to SMD on educational attainment

We found that genetic liability to schizophrenia had a small positive effect on EA only after including income in the model (MVMR-IVMSCZ=1.046 months per doubling of genetic liability to schizophrenia, P-value= 0.0000057), while genetic liability to BD had significant total and direct effects on EA (MVMR-IVMBIP=1.254 months, P-value= 0.00817). Genetic liability to MDD had a small negative total effect on EA in the univariate analyses (IVWMDD=-2.79 months, P-value=0.0211) that was no longer significant after accounting for income (see eTable1,eTable4 and Figure 1C).

#### Effects of genetic liability to SMD on household income

Genetic liability to schizophrenia had significant total and direct negative effects on household income (IVWSCZ=-819.99£, P-value=7.55E-08, MVMR-IVMSCZ=-960.41£, P-value=5.27E-09). We also found that genetic liability to depression had a significant negative total effect on income (IVWDEP=-2639£, P-value=8.69e-04), with comparable albeit slightly lower direct effects after accounting for EA in the MVMR models. There was no significant effect of genetic liability to BD on income (see Table1,eTable5 and Figure1D)

## Direct effects of genetic liability to socioeconomic status and intelligence on the risk of SMD

Considering previous inconsistent evidence of a protective effect of education on mental disorders,^48^ we conducted supplementary post-hoc MVMR analyses to assess the potential role of intelligence in the positive association found between genetic liability to EA and the risk of schizophrenia and BD. We found that the direct effect of EA on the risk of BD and schizophrenia, not mediated through intelligence, was larger than that found in the original model (MVMR-OR_SCZ_=3.19 per-SD-increase, P-value=5.38E-07; IVW-ORBIP=3.60, P-value= 3.06E-07). There was a direct negative effect of intelligence, not mediated by EA, with around 0.35-fold decreased risk per-SD-increase on schizophrenia and BD (MVMR-OR_SCZ_=0.66, P-value=0.0054; MVMR-OR_BIP_=0.67, P-value: 0.0119). Income effects were practically unchanged. Our results suggested that genetic liability to intelligence, not mediated by EA, was associated with increased risk of depression (MVMR-ORDEP=1.25 per-SD-increase, P-value=3.03E-05) (see Figure 2 and eTable6).

**Figure 2:**
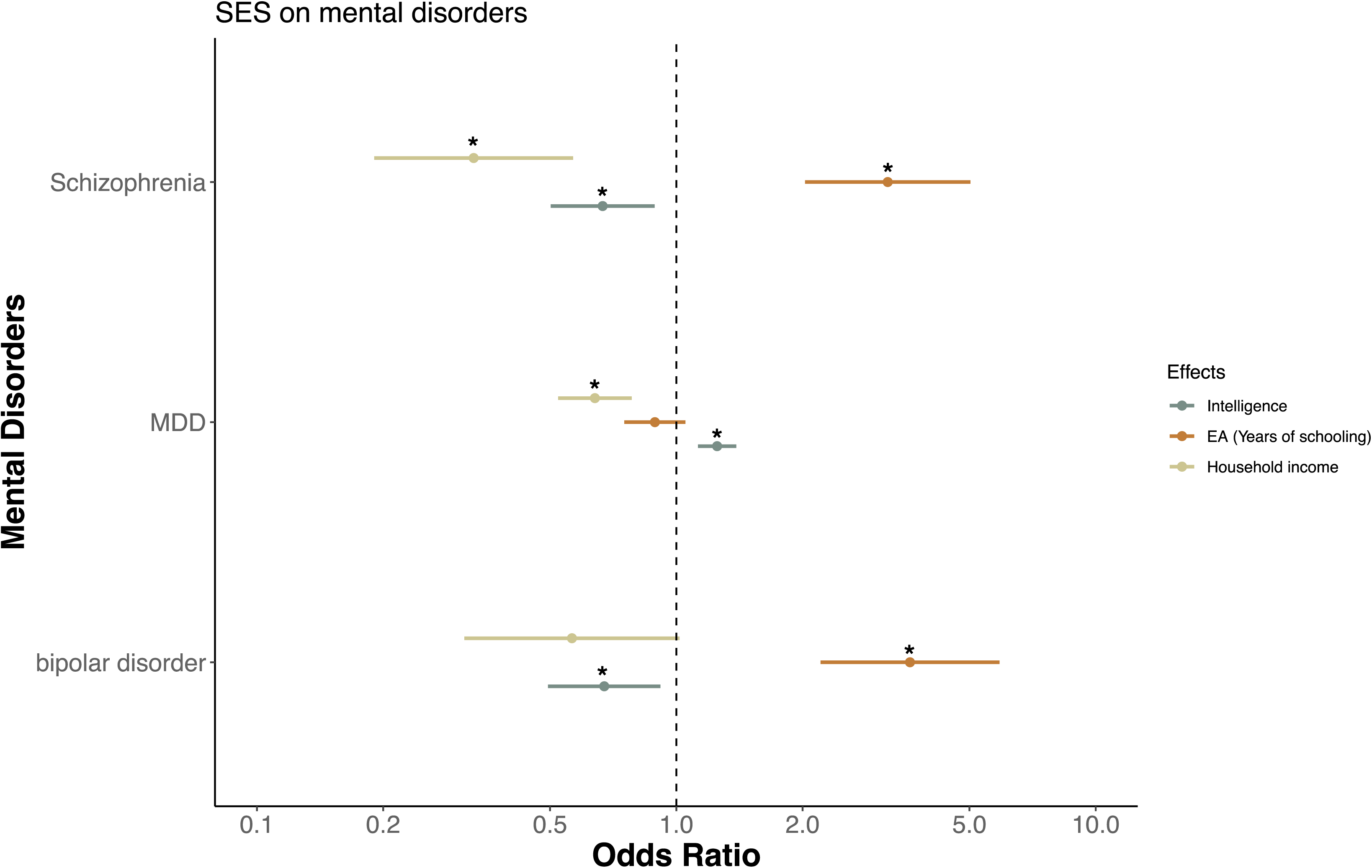
Direct effects of genetic liability to intelligence and SES on the risk of SMD: Follow-up multivariable Mendelian randomisation analyses of genetic liability to intelligence, EA, and income on the risk of each SMD. 95% confidence intervals included after non-parametric 1,000 iteration bootstrap permutations are marked with an asterisk

## DISCUSSION

Despite the well-established relationship of socioeconomic status (SES) to mental disorders,^17,18^ considerable debate remains about the causal links between key SES constituents income and education with SMD.^17,49,50^ We used MVMR to address the potential influence of SES multidimensionality on mental disorders by analysing the independent effects of genetic liability to income and EA on three SMD. Our study revealed different patterns of bidirectional associations between SES constituents and the three SMD. We observed a negative bidirectional association between household income on the risk of schizophrenia and MDD, with a smaller reverse causal effect. On the contrary, we found a positive bidirectional relationship of genetic liability to EA with schizophrenia and BD which only becomes apparent after adjusting for income. These findings are in line with a recent nationwide study in Finland supporting contributions of both social causation and health selection mechanisms to the risk of mental disorders^50^, and may contradict, at least in terms of genetic liability, widespread social causation vs health selection theories.^51^

EA MVMR estimates were in agreement with previous positive genetic correlations with schizophrenia and bipolar disorder.^52^ However, as schizophrenia has been classically associated with premorbid cognitive deficits and cognitive decline,^53,54^ we conducted post-hoc analyses including genetic liability to intelligence in the MVMR model to assess these apparently counter-intuitive results. We found an even greater positive direct effect of EA (independently of intelligence and income, and thus probably reflecting non-cognitive skills^55^) on the risk of bipolar disorder and schizophrenia. By contrast, we found that, after adjusting for EA and income, intelligence was associated with decreased risk for both schizophrenia and BD. These results are consistent with genetic correlations of BD and schizophrenia with cognitive and non-cognitive skills latent factors shown by the recent GWAS-by-subtraction method^56^ and also support findings from genetic studies showing a polygenic contribution of schizophrenia and BD liability to creative and artistic careers^57^ as opposed to the negative association of genetic liability to both disorders with general cognitive function.^33,58^ Our results suggest that non-cognitive skills such as creativity, higher tolerance of risk, or perseverance towards an activity, which may provide evolutionary benefits, could also lead to an increased susceptibility to schizophrenia or BD.^57,59^

Overall, our research supports previous evidence linking socioeconomic inequality and poor mental health,^7,17,60^ but at the same time highlights the caution to be adopted when assessing socio-environmental phenomena through genetic data. In the United States, the estimated economic burden of schizophrenia was around $150 billion per year, mainly due to indirect costs associated with unemployment and lack of productivity.^61^ Our results support an association between genetic liability to schizophrenia and MDD and lower household income, but the effects ware smaller than those expected from the observational data. MR analyses with dichotomized binary (i.e., mental disorders) or categorical ordinal exposures (i.e., income or EA) capture genetic liability to an exposure rather than pure variation in the exposure category, which may impact the magnitude of the effects and in particular of socioeconomic traits.^47^ A combined liability-threshold model which includes putative rare variants with large effects^62^ and involves potential gene-environmental interaction effects (GxE) such as the stigma and discrimination associated with some diagnostic categories,^63^ the disability associated with early symptoms and drug abuse,^64,65^ the potential impact of isolation and social defeat^66,67^ and a likely negative expectation effect towards SMD such as schizophrenia,^68^ could provide more accurate estimates in the future.

Our findings are also consistent with previous genome-wide association studies showing that income-related genetic variants are more strongly associated with better mental health than education genetic variables.^23^ We found that the total effects of genetic liability to EA on the three mental disorders tended to diverge from direct effects after accounting for income. For instance, we only found a positive bidirectional connection between schizophrenia and EA after adjusting for income, while the bidirectional negative association of MDD with EA was only significant in the unadjusted analyses and was thus probably confounded by income. On the contrary, the pattern of bidirectional associations between household income and SMD was less influenced by EA overall. Our results suggest that we should assess the independent and joint effects of income and EA to better address the complexity of their causal links to SMD. They also underscore the role of novel multivariate methods such as MVMR in disentangling the effects of complex phenotypes with highly correlated sub-phenotypes such as SES.

### Strengths and limitations

Our study has several strengths. First, we avoided the effects of common unmeasured confounding factors found in classical observational studies and reverse causality by using MR. The large sample size of the available GWAS provided adequate statistical power for causal inferences and we minimized the potential exposure-outcome bias by restricting our main results to non-overlapping samples. We also performed a large number of sensitivity analyses in the presence of weak instruments.^32^ However, our study also has several limitations. First, recent studies have highlighted that MR analyses and SES phenotypes may be affected by assortative mating, dynastic effects, participation bias and design assumptions.^48,69–72^ Second, our study does not allow us to completely disentangle the potential influence of parental SES on the risk of severe mental illness^28^ and our findings should only be interpreted in terms of genetic liability in epidemiology.^47^ In the UK biobank sample, income was measured at the household level. Previous studies have shown that there is a genetic overlap between household income levels and individual income, as well as a high correlation with individual EA^23,73^ thus minimizing these potential biases. Third, there is a shared genetic contribution between mental disorders and socioeconomic factors.^26^ We found a widespread heterogeneity in our analyses and the genetic variants used as instrumental variables could have been biased by pleiotropic effects and conditional instrumental strength. We tried to address these limitations by using MVMR robust-to-weak instruments methods. Future research may benefit from new approaches like within-family MR designs^74^ or negative control methods^69^ that are currently being developed to reduce the likelihood of these biases and limitations. Finally, our study is restricted to the European population, which limits the generalizability of our results to other locations and populations. The impact of socioeconomic determinants on mental health varies across contexts, as exemplified by divergent effects of EA and income in studies in Latin American and British populations.^75^ More diversity of genetic samples and research across a broader range of regions and populations, including participants from low-or middle-income countries, would contribute to a more comprehensive understanding of the complex relationships between socioeconomic factors and global mental health.

## Conclusions

We provide evidence that genetically predicted income was associated with a lower risk of schizophrenia and MDD and that schizophrenia and MDD were associated with decreased income. On the contrary, a positive bidirectional association of genetic liability to EA with schizophrenia and BD was found, but only after adjusting for income. Our study advances knowledge of the mechanisms by which social determinants influence mental health by dissecting the independent effects of income and EA on different SMD and may help to guide public health strategies for allocating mental health resources and addressing social inequality.

## Supporting information

eMethods and eTables

## Data Availability

All data produced are available online at https://www.ukbiobank.ac.uk/https://pgc.unc.edu/https://gwas.mrcieu.ac.uk/

## Acknowledgements

This work was supported by the Spanish Ministry of Science and Innovation. Instituto de Salud Carlos III (SAM16PE07CP1, PI16/02012, PI17/00997, PI19/01024, PI20/00721), co-financed by ERDF Funds from the European Commission, “A way of making Europe”, CIBERSAM. Madrid Regional Government (B2017/BMD-3740 AGES-CM-2), European Union Structural Funds. European Union Seventh Framework Program under grant agreements FP7-4-HEALTH-2009-2.2.1-2-241909 (Project EU-GEI), FP7-HEALTH-2013-2.2.1-2-603196 (Project PSYSCAN), and FP7-HEALTH-2013-2.2.1-2-602478 (Project METSY); and European Union H2020 Program under the Innovative Medicines Initiative 2 Joint Undertaking (grant agreement No 115916, Project PRISM, and grant agreement No 777394, Project AIMS-2-TRIALS), Fundación Familia Alonso, Fundación Alicia Koplowitz, and Fundación Mutua Madrileña. A.A.-B. holded a Rio Hortega Grant during the development of the research from Instituto de Salud Carlos III (CM20/00114). C.M.D.-C. holds a Juan Rodés Grant from Instituto de Salud Carlos III (JR19/00024).

## Ethics declarations

### Competing interests

C.A has been a consultant to or has received honoraria or grants from Acadia, Angelini, Gedeon Richter, Janssen Cilag, Lundbeck, Minerva, Otsuka, Roche, Sage, Servier, Shire, Schering Plough, Sumitomo Dainippon Pharma, Sunovion, and Takeda. C.M.D.-C. has received honoraria from AbbVie, Sanofi, and Exeltis. The rest of the authors declare no competing interests.

