## Supplementary material for "Socioeconomic status and severe mental disorders: A Bidirectional Multivariable Mendelian Randomisation Study": eMethods and eTables

### Supplemental Online Content

**STROBE-MR checklist**

**eMethods** Detailed Methods.

**eTable 1.** Univariable and Multivariable mendelian randomisation results of socioeconomic status phenotypes on mental disorders**.**

**eTable 2.** Univariable Mendelian Randomisation Results and sensitivity analyses of genetic liability to Educational attainment Versus Severe mental Disorders

**eTable 3.** Univariable Mendelian Randomisation Results and sensitivity analyses of genetic liability to household income Versus Severe mental Disorders

**eTable 4.** Univariable Mendelian Randomisation Results and sensitivity analyses of genetic liability to severe mental disorders Versus Educational attainment

**eTable 5.** Univariable Mendelian Randomisation Results and sensitivity analyses of genetic liability to severe mental disorders Versus household income

**eTable 6.** Multivariable Mendelian Randomisation Results of Socioeconomic status and Intelligence versus severe mental disorders

#### STROBE-MR checklist of recommended items to address in reports of Mendelian randomization studies1, 2

| Item No. | Section | Checklist item | Page No. | Relevant text from manuscript |
|---|---|---|---|---|
| 1 | **TITLE and ABSTRACT** | Indicate Mendelian randomization (MR) as the study's design in the title and/or the abstract if that is a main purpose of the study | 1 | Socioeconomic status and severe mental disorders: A Bidirectional Multivariable Mendelian Randomisation Study |
| | **INTRODUCTION** | | | |
| 2 | **Background** | Explain the scientific background and rationale for the reported study. What is the exposure? Is a potential causal relationship between exposure and outcome plausible? Justify why MR is a helpful method to address the study question | 2-3 | "Extensive evidence based on survey data, large cohort observational studies and family-level studies supports the association between mental disorders and lower income, disadvantaged socioeconomic position and homelessness" […] "However, the specific pattern of relationships is heterogeneous, as some disorders such as ASD or bipolar disorder (BD) are associated with higher parental or individual socioeconomic status,9,10 while depression or schizophrenia are associated with lower levels of education and income"<br>"Despite the efforts to disentangle the complex relationship between SES and mental disorders, there is still ongoing debate due to the disparity of results and lack of replication.8,17,28 Further, conventional observational studies are affected by unmeasured confounders that preclude adequate assessment of the directional associations between SES and mental disorders.11 The use of univariable Mendelian randomisation (UVMR) has become popular for analysing causality between exposures to risk or protective factors and outcomes such as diseases in recent years, to overcome the limitations of observational studies.29,30 An extension of MR called multivariable Mendelian randomisation (MVMR) has recently been developed to allow the analysis of the independent causal effects of correlated exposures simultaneously. |
| 3 | **Objectives** | State specific objectives clearly, including pre-specified causal hypotheses (if any). State that MR is a method that, under specific assumptions, intends to estimate causal effects | 3 | We performed bidirectional two-sample MR to determine whether genetic liability to household income and educational attainment were causally linked to three SMD (i.e., schizophrenia (SCZ), bipolar disorder (BD) and major depressive disorder (MDD)) with onset in adolescence or young adulthood and differing effects across the psychotic, cognitive and affective spectra. We used MVMR to disentangle the independent contribution of income and EA to SMD and the independent reverse effects of SMD on each SES trait. |
| | **METHODS** | | | |

| | | | | |
|---|---|---|---|---|
| 4 | **Study design and data sources** | Present key elements of the study design early in the article. Consider including a table listing sources of data for all phases of the study. For each data source contributing to the analysis, describe the following: | | |
| | a) | Setting: Describe the study design and the underlying population, if possible. Describe the setting, locations, and relevant dates, including periods of recruitment, exposure, follow-up, and data collection, when available. | 4 | We performed bidirectional two-sample UVMR and MVMR analyses to assess the causal links between SES (income and EA) and SMD in public datasets of European population. We performed follow-up analyses including genetic liability to intelligence33 in MVMR models to assess the causal links independently of intelligence |
| | b) | Participants: Give the eligibility criteria, and the sources and methods of selection of participants. Report the sample size, and whether any power or sample size calculations were carried out prior to the main analysis | 4 | We used the largest publicly available genome-wide association study (GWAS) summary statistics of household income from the UK Biobank(n= 397,751)35. For EA we used the largest available GWAS from European Ancestry (n=766,345)24. The EA phenotype was a meta-analysis of 70 GWAS with standardised measures of years of schooling, including 442,183 individuals from the UK Biobank cohort.<br><br>Mental Disorders<br>We selected the latest GWAS from the Psychiatric Genomics Consortium (PGC) for (i) SCZ (Wave-3, only European ancestry,36 comprising 52,017 cases and 75,889 controls); (ii) BD37, 20,352 cases and 31,358 controls; and (iii) MDD38, 170,756 cases and 329,443 controls.<br><br>Intelligence<br>We used the largest GWAS meta-analysis of intelligence comprising 14 independent epidemiological cohorts of European ancestry (n= 269,867). |
| | c) | Describe measurement, quality control and selection of genetic variants | | Not applicable |
| | d) | For each exposure, outcome, and other relevant variables, describe methods of assessment and diagnostic criteria for diseases | | Not applicable |
| | e) | Provide details of ethics committee approval and participant informed consent, if relevant | 4 | The ethics declarations for each public dataset used in the present research can be found in their original publications. |

| | | | | | |
|---|---|---|---|---|---|
| 5 | **Assumptions** | | Explicitly state the three core IV assumptions for the main analysis (relevance, independence and exclusion restriction) as well assumptions for any additional or sensitivity analysis | eMethods | For both univariable(UVMR) and multivariable (MVMR) analyses, IVs must satisfy the following three main assumptions: (i) relevance (i.e. variants are robustly associated with the exposures (related to weak instrument bias); (ii) exchangeability n (i.e. variants are independent of confounders of both exposures and outcomes); and (iii) exclusive restriction (i.e. the relationship of the SNPs with the outcome is only due to their relationship with the exposure). We performed a bidirectional univariable and multivariable two-sample Mendelian Randomization (UVMR and MVMR) to assess the causal link between socioeconomic status(income and EA) and severe mental disorders. We performed post-hoc analyses including the cognitive ability phenotype in MVMR models to assess causal directions independently of intelligence. |
| 6 | **Statistical methods: main analysis** | | Describe statistical methods and statistics used | 5 | |
| | | a) | Describe how quantitative variables were handled in the analyses (i.e., scale, units, model) | | Not applicable |
| | | b) | Describe how genetic variants were handled in the analyses and, if applicable, how their weights were selected | | We extracted genome-wide significant SNPs as instrumental variables at p < 5 × 10−8. |
| | | c) | Describe the MR estimator (e.g. two-stage least squares, Wald ratio) and related statistics. Detail the included covariates and, in case of two-sample MR, whether the same covariate set was used for adjustment in the two samples | | We used an inverse-variance weighted method (IVW) as the main analysis, as recommended by the guidelines due to its robustness.<br>The total effects provided by a classical UVMR method can suffer from bias when there is a high correlation between exposures, as is the case of income and EA. Hence, we evaluated i) the direct effect independent of EA of genetic liability to income on each SMD, and the direct effect of genetic liability to each SMD on income and ii) the direct effect independent of income of genetic liability to EA on each SMD, and the direct effect of genetic liability to each SMD on EA |
| | | d) | Explain how missing data were addressed | | Not applicalbe |
| | | e) | If applicable, indicate how multiple testing was addressed | | |
| 7 | **Assessment of assumptions** | | Describe any methods or prior knowledge used to assess the assumptions or justify their validity | 5 | For each exposure, we calculated the F statistic to analyse the instrument's strength39 and used the rule of thumb of F>10 adopted in previous studies to define weak instrument bias” |

| | | | | | |
|---|---|---|---|---|---|
| | | | | | We estimated the instrumental variable strength using conditional-F-statistics and heterogeneity as well as horizontal pleiotropy using a modified Cochran's Q-statistic. |
| 8 | **Sensitivity analyses and additional analyses** | | Describe any sensitivity analyses or additional analyses performed (e.g. comparison of effect estimates from different approaches, independent replication, bias analytic techniques, validation of instruments, simulations) | | we used Weighted median (WM)14, MR-Egger15, Simple mode (SM), Weighted mode (WMo) heterogeneity tests (IVW, and Egger Cochran's Q statistic test), and horizontal pleiotropy tests (Egger intercept) as sensitivity analyses15. Additionally, we conducted Mendelian Randomization Pleiotropy RESidual Sum and Outlier (MR-PRESSO)<br>heterogeneity as well as horizontal pleiotropy using a modified Cochran's Q-statistic. We performed IVM-MVMR regression of each model and robust-to-weak-instrument Q-statistic minimization, using non-parametric 1000 iteration bootstrap permutations |
| 9 | **Software and pre-registration** | | | | |
| | | a) | Name statistical software and package(s), including version and settings used | 5 | All analyses were performed using the "TwoSampleMR" package v 0.5.6, with R version 4.1.2<br>MVMR analyses were performed with the MVMR package (more details in eMethod and [https://github.com/WSpiller/MVMR]) |
| | | b) | State whether the study protocol and details were pre-registered (as well as when and where) | | Not applicable |
| | **RESULTS** | | | | |
| 10 | **Descriptive data** | | | | |
| | | a) | Report the numbers of individuals at each stage of included studies and reasons for exclusion. Consider use of a flow diagram | | Not applicable |
| | | b) | Report summary statistics for phenotypic exposure(s), outcome(s), and other relevant variables (e.g. means, SDs, proportions) | | Analyses exploring the effect of SMD on EA and income were first multiplied by the SD of income(SD= 33,181£)44 and EA(SD=4.2 years of education)24 to convert them to British pounds and months and then expressed as the effect of a doubling (2-fold increase) of genetic liability to each SMD on the odds of income and EA, as recommended in previous studies |

| | | | | | |
|---|---|---|---|---|---|
| | | c) | If the data sources include meta-analyses of previous studies, provide the assessments of heterogeneity across these studies | eMethods | Despite sample and methodological differences, genetic correlations (rg) between cohorts were substantial (mean=0.67), and there was no evidence of cohort heterogeneity in SNP associations. |
| | | d) | For two-sample MR:<br>i. Provide justification of the similarity of the genetic variant-exposure associations between the exposure and outcome samples<br>ii. Provide information on the number of individuals who overlap between the exposure and outcome studies | eMethods | Following the recommendations from the authors of MVMR, we estimated the covariances of the model including EA and income using the genetic correlation between both phenotypes in the UK biobank (rg= 0.87)4 as a proxy of their phenotypic covariance, considering an almost complete overlap between the samples. For the post-hoc intelligence analyses we used the same proxy phenotypic covariance with a matrix with the genetic correlations(rg) between income, educational attainment and intelligence:<br><br>1 1.00 0.63 0.74<br>2 0.74 1.00 0.87<br>3 0.63 0.87 1.00<br><br>For mental disorders, the covariances between SES phenotypes and SMD were 0 by design, as there was no overlap between samples.<br><br>There is no overlap between the individuals in the SES phenotype datasets and the SMD datasets |
| 11 | **Main results** | | | 6-7 | |
| | | a) | Report the associations between genetic variant and exposure, and between genetic variant and outcome, preferably on an interpretable scale | | Not applicable |
| | | b) | Report MR estimates of the relationship between exposure and outcome, and the measures of uncertainty from the MR analysis, on an interpretable scale, such as odds ratio or relative risk per SD difference | 6-7<br>eTable1 | MVMR-IVMSCZ=1.046 months per doubling of genetic liability to schizophrenia, P-value= 0.0000057), while genetic liability to BD had significant total and direct effects on EA (MVMR-IVMBIP=1.254 months, P-value= 0.00817). Genetic liability to MDD had a small negative total effect on EA in the univariate analyses (IVWMDD=-2.79 months, P-value=0.0211) |

| | | | | | |
|---|---|---|---|---|---|
| | | | | | IVWSCZ=-819.99£, P-value=7.55E-08, MVMR-IVMSCZ=-960.41£, P-value=5.27E-09). We also found that genetic liability to depression had a significant negative total effect on income (IVWDEP=-2639£, P-value=8.69e-04), with comparable albeit slightly lower direct effects after accounting for EA in the MVMR models. There was no significant effect of genetic liability to BD on income<br>[…] |
| | | c) | If relevant, consider translating estimates of relative risk into absolute risk for a meaningful time period | | Not applicable |
| | | d) | Consider plots to visualize results (e.g. forest plot, scatterplot of associations between genetic variants and outcome versus between genetic variants and exposure) | Figure 1 and 2 | |
| 12 | **Assessment of assumptions** | | | | |
| | | a) | Report the assessment of the validity of the assumptions | eMethods | Sensitivity analyses mirrored the results of the IVW method and egger-intercept showed non-significant results indicating that there was little evidence for directional genetic pleiotropy(eTable 2A,2B<br>We found heterogeneity for both disorders, but there was limited evidence of horizontal pleiotropy, as suggested by the MR-Egger intercept (see eTable1 and 3) |
| | | b) | Report any additional statistics (e.g., assessments of heterogeneity across genetic variants, such as $I^2$, Q statistic or E-value) | | eTable 1,2,3,4,5,6 |
| 13 | **Sensitivity analyses and additional analyses** | | | eTable 1,2,3,4 | eTable 1,2,3,4,5,6 |
| | | a) | Report any sensitivity analyses to assess the robustness of the main results to violations of the assumptions | | |
| | | b) | Report results from other sensitivity analyses or additional analyses | | |

| | | | | | |
|---|---|---|---|---|---|
| | | c) | Report any assessment of direction of causal relationship (e.g., bidirectional MR) | | |
| | | d) | When relevant, report and compare with estimates from non-MR analyses | | |
| | | e) | Consider additional plots to visualize results (e.g., leave-one-out analyses) | | |
| | **DISCUSSION** | | | | |
| 14 | **Key results** | | Summarize key results with reference to study objectives | 8 | Our study revealed different patterns of bidirectional associations between SES constituents and the three SMD. We observed a negative bidirectional association between household income on the risk of schizophrenia and MDD, with a smaller reverse causal effect. On the contrary, we found a positive bidirectional relationship of genetic liability to EA with schizophrenia and BD which only becomes apparent after adjusting for income. |
| 15 | **Limitations** | | Discuss limitations of the study, taking into account the validity of the IV assumptions, other sources of potential bias, and imprecision. Discuss both direction and magnitude of any potential bias and any efforts to address them | 9 | However, our study also has several limitations. First, recent studies have highlighted that MR analyses and SES phenotypes may be affected by assortative mating, dynastic effects, participation bias and design assumptions.48,69–72 Second, our study does not allow us to completely disentangle the potential influence of parental SES on the risk of severe mental illness28 and our findings should only be interpreted in terms of genetic liability in epidemiology.47 In the UK biobank sample, income was measured at the household level. Previous studies have shown that there is a genetic overlap between household income levels and individual income, as well as a high correlation with individual EA23,73 thus minimizing these potential biases. Third, there is a shared genetic contribution between mental disorders and socioeconomic factors.26 We found a widespread heterogeneity in our analyses and the genetic variants used as instrumental variables could have been biased by pleiotropic effects and conditional instrumental strength. We tried to address these limitations by using MVMR robust-to-weak instruments methods. Future research may benefit from new approaches like within-family MR designs74 or negative control methods69 that are currently being developed to reduce the likelihood of these biases and limitations. Finally, our study is restricted to the European population, which limits the generalizability of our results to other locations and populations. The impact of socioeconomic determinants on mental health varies across contexts, as exemplified by divergent effects of EA and income in studies in Latin American and British populations |
| 16 | **Interpretation** | | | | |
| | | a) | Meaning: Give a cautious overall interpretation of results in the context of their | 9 | We provide evidence that genetically predicted income was associated with a lower risk of schizophrenia and MDD and that schizophrenia and MDD were associated with decreased income. On the contrary, a positive |

| | | | | | |
|---|---|---|---|---|---|
| | | | limitations and in comparison with other studies | | bidirectional association of genetic liability to EA with schizophrenia and BD was found, but only after adjusting for income. |
| | | b) | Mechanism: Discuss underlying biological mechanisms that could drive a potential causal relationship between the investigated exposure and the outcome, and whether the gene-environment equivalence assumption is reasonable. Use causal language carefully, clarifying that IV estimates may provide causal effects only under certain assumptions | | Not applicable |
| | | c) | Clinical relevance: Discuss whether the results have clinical or public policy relevance, and to what extent they inform effect sizes of possible interventions | | Our study advances knowledge of the mechanisms by which social determinants influence mental health by dissecting the independent effects of income and EA on different SMD and may help to guide public health strategies for allocating mental health resources and addressing social inequality |
| 17 | **Generalizability** | | Discuss the generalizability of the study results (a) to other populations, (b) across other exposure periods/timings, and (c) across other levels of exposure | | The impact of socioeconomic determinants on mental health varies across contexts, as exemplified by divergent effects of EA and income in studies in Latin American and British populations.75 More diversity of genetic samples and research across a broader range of regions and populations, including participants from low- or middle-income countries, would contribute to a more comprehensive understanding of the complex relationships between socioeconomic factors and global mental health |
| | **OTHER INFORMATION** | | | | |
| 18 | **Funding** | | Describe sources of funding and the role of funders in the present study and, if applicable, sources of funding for the databases and original study or studies on which the present study is based | 19 | Acknowledgements<br>This work was supported by the Spanish Ministry of Science and Innovation. Instituto de Salud Carlos III (SAM16PE07CP1, PI16/02012, PI17/00997, PI19/01024, PI20/00721), co-financed by ERDF Funds from the European Commission, “A way of making Europe”, CIBERSAM. Madrid Regional Government (B2017/BMD-3740 AGES-CM-2), European Union Structural Funds. European Union Seventh Framework Program under grant agreements FP7-4-HEALTH-2009-2.2.1-2-241909 (Project EU-GEI), FP7- HEALTH-2013-2.2.1-2-603196 (Project PSYSCAN), and FP7- HEALTH-2013-2.2.1-2-602478 (Project METSY); and European Union H2020 Program under the Innovative Medicines Initiative 2 Joint Undertaking (grant agreement No 115916, Project PRISM, and grant agreement No 777394, Project AIMS-2-TRIALS), Fundación Familia Alonso, Fundación Alicia Koplowitz, and Fundación Mutua Madrileña. A.A.-B. holded a Rio Hortega Grant during the development of the research from Instituto de Salud Carlos III (CM20/00114). C.M.D.-C. holds a Juan Rodés Grant from Instituto de Salud Carlos III (JR19/00024). |

| | | | | |
|---|---|---|---|---|
| 19 | **Data and data sharing** | Provide the data used to perform all analyses or report where and how the data can be accessed, and reference these sources in the article. Provide the statistical code needed to reproduce the results in the article, or report whether the code is publicly accessible and if so, where | | We used the largest publicly available genome-wide association study (GWAS) summary statistics of household income from the UK Biobank<br>We selected the latest GWAS from the Psychiatric Genomics Consortium (PGC) |
| 20 | **Conflicts of Interest** | All authors should declare all potential conflicts of interest | 19 | C.A has been a consultant to or has received honoraria or grants from Acadia, Angelini, Gedeon Richter, Janssen Cilag, Lundbeck, Minerva, Otsuka, Roche, Sage, Servier, Shire, Schering Plough, Sumitomo Dainippon Pharma, Sunovion, and Takeda. C.M.D.-C. has received honoraria from AbbVie, Sanofi, and Exeltis. The rest of the authors declare no competing interests. |

This checklist is copyrighted by the Equator Network under the Creative Commons Attribution 3.0 Unported (CC BY 3.0) license.

1. Skrivankova VW, Richmond RC, Woolf BAR, Yarmolinsky J, Davies NM, Swanson SA, et al. Strengthening the Reporting of Observational Studies in Epidemiology using Mendelian Randomization (STROBE-MR) Statement. JAMA. 2021;under review.

2. Skrivankova VW, Richmond RC, Woolf BAR, Davies NM, Swanson SA, VanderWeele TJ, et al. Strengthening the Reporting of Observational Studies in Epidemiology using Mendelian Randomisation (STROBE-MR): Explanation and Elaboration. BMJ. 2021;375:n2233.

**eMethods: Detailed Methodology.**

**Study design**

Mendelian Randomization (MR) is an instrumental variable (IV) analysis method based on genetic variants (Single nucleotide polymorphisms, SNPs). MR enables causal estimates between exposure and an outcome thanks to the natural randomization of genetic variants that takes place in meiosis[1]. MVMR is an extension of the MR method that allows the inclusion of IVs of different exposures that are phenotypically and genetically associated in the same model in order to obtain the independent direct effects of each exposure on the outcome[2,3].

**Mendelian randomization assumptions:**

For both univariable(UVMR) and multivariable (MVMR) analyses, IVs must satisfy the following three main assumptions: (i) relevance (i.e. variants are robustly associated with the exposures (related to weak instrument bias); (ii) exchangeability n (i.e. variants are independent of confounders of both exposures and outcomes); and (iii) exclusive restriction (i.e. the relationship of the SNPs with the outcome is only due to their relationship with the exposure). We performed a bidirectional univariable and multivariable two-sample Mendelian Randomization (UVMR and MVMR) to assess the causal link between socioeconomic status(income and EA) and severe mental disorders. We performed post-hoc analyses including the cognitive ability phenotype in MVMR models to assess causal directions independently of intelligence.

**Additional information of Data sources and Genetic instruments**

Socioeconomic Status Phenotypes

We used the latest publicly available genetic summary statistics GWAS of household income [4] from the MRC-IEU UK Biobank GWAS Pipeline (n= 397,751) estimated from the household income Phesant-derived phenotype which comprised five-category ordinal midpoints from "less than 18,000£" up to "more than 100,000£" with an estimated mean income of 44,409£ (standard deviation: 33,181£)[5].

For Educational Attaniment we used the largest available genetic summary statistics from European Ancestry, excluding 23andMe individuals (n= 766,345)[6] and including 442,183 individuals from the UK Biobank cohort. The EA phenotype was a meta-analysis of 70 GWAS with standardized measures of years of schooling (weighted mean 16.8 years, standard deviation 4.2).

Mental Disorders

We selected summary statistics GWAS from the Psychiatric Genetic Consortium (PGC) for (i) Schizophrenia ( SCZ PGC Wave 3) we used only European ancestry summary statistics,[7]comprising 52,017 cases and 75,889 controls. (ii) Bipolar disorder (BD)[8], comprising 20,352 cases and 31,358 controls; and (iii) Major depressive disorder (MDD)[9], comprising 170,756 cases and 329,443 controls. There is no overlap between the individuals in the SES phenotype datasets and the SMD datasets.

<u>Intelligence phenotype</u>
We used the largest GWAS meta-analysis of intelligence comprising 14 independent epidemiological cohorts of European ancestry (n= 269,867).[10] The selected studies employed a comprehensive approach to evaluate intelligence through the utilization of various neurocognitive tests, with a primary focus on assessing the dynamic components of cognitive abilities. Despite variations in the specific formats and subject matter of these tests, the resulting scores consistently demonstrated a robust positive correlation pattern, establishing a dependable empirical observation that extends across diverse populations. From a statistical perspective, the shared variability among cognitive tasks can be effectively represented as an underlying factor known as "g-factor", denoting the general factor of intelligence. Despite sample and methodological differences, genetic correlations (rg) between cohorts were substantial (mean=0.67), and there was no evidence of cohort heterogeneity in SNP associations.

We extracted genome-wide significant SNPs as instrumental variables (IV) at $p < 5 \times 10{-}8$. For every GWAS dataset, we applied a LD-Clumping R2 threshold of 0.001 and a window of 10,000 kb based on the European phase 3 reference panel for each phenotype. Exposure and outcome sets were harmonized and palindromic SNPs with intermediate allele frequencies were excluded. For each exposure, we calculated the F statistic to analyse the instrument's strength[11] and used the rule of thumb of F>10 adopted in previous studies to define weak instrument bias[12,13].

All studies received ethical approval from their respective institutional review committees, including informed consent from participants and rigorous quality control that can be found in the original publications.

**Statistical analysis**

<u>Univariable Mendelian Randomization</u>
We conducted a bidirectional two-sample UVMR analysis to assess putative causality between household income and EA and the three SMD. We used an inverse-variance weighted method (IVW) as the main analysis, as recommended by the guidelines due to its robustness[12]. However, as

IVW may generate biased results in case of heterogeneity or violation of any assumption, we used Weighted median (WM)[14], MR-Egger[15], Simple mode (SM), Weighted mode (WMo)[16], heterogeneity tests (IVW, and Egger Cochran's Q statistic test), and horizontal pleiotropy tests (Egger intercept) as sensitivity analyses[15]. Additionally, we conducted Mendelian Randomization Pleiotropy RESidual Sum and Outlier (MR-PRESSO)[17] which allows correction of horizontal pleiotropy via outlier removal. For MR-PRESSO we used NbDistribution = 2000, SignifThreshold = 0.05 as regular parameters. All analyses were performed using the "TwoSampleMR" package v 0.5.6 [11], R Version 1.2.1335.

Multivariable Mendelian Randomization

Multivariable Mendelian randomization (MVMR) is a newly developed method to estimate the direct effects of correlated exposures using the overlapping set of instruments in the same model. The total effect provided by a classical MR method could suffer from bias when there is a high correlation between exposures, as in the case of income and educational attainment. We evaluated i) the direct effect (i.e. independent of EA) of genetic liability to income on mental disorders, and the direct effect of genetic liability to Schizophrenia, Bipolar, and MDD on income and ii) the direct effect (i.e. independent of income) of genetic liability to EA on mental disorders, and the direct effect of genetic liability to mental disorders, on EA. Finally, we conducted supplementary post-hoc analyses including the cognitive ability phenotype in the MVMR models to assess the association of the genetic liability of EA and income with SMD independently of intelligence.

For each MVMR analysis, we applied the same LD-Clumping R2 threshold (0.001 and a window of 10,000 kb) using the "TwosampleMR" package[11]. Then, the exposure and outcome data were harmonized by aligning the effect alleles of SNP to the human genome reference sequence using the IEU GWAS database as for UVMR, ensuring that the effect of each SNP on the exposure and outcome corresponded to the same allele. We then followed the recent pipeline developed by Sanderson et al (2021)[2] to estimate causal effects and correct for potential weak and pleiotropic instruments through various sensitivity analyses. Following the recommendations from the authors of MVMR, , we estimated the covariances of the model including EA and income using the genetic correlation between both phenotypes in the UK biobank (rg= 0.87)[4] as a proxy of their phenotypic covariance, considering an almost complete overlap between the samples. For the post-hoc intelligence analyses we used the same proxy phenotypic covariance with a matrix with the genetic correlations(rg) between income, educational attainment and intelligence:

| | | | |
|---|---|---|---|
| **1** | 1.00 | 0.63 | 0.74 |
| **2** | 0.74 | 1.00 | 0.87 |
| **3** | 0.63 | 0.87 | 1.00 |

For mental disorders, the covariances between SES phenotypes and SMD were 0 by design, as there was no overlap between samples.

We estimated the strength of instruments using conditional F statistics and heterogeneity as well as horizontal pleiotropy using a modified Cochran's Q statistic across the set of instruments. We performed inverse variance weighted (IVW) MVMR regression of each model and robust to weak instrument MVMR through Q statistic minimization using non-parametric 1000 iteration bootstrap permutations.

All MVMR analyses were performed with the MVMR package, for details on the method and work pipeline see the original paper[2] and [https://github.com/WSpiller/MVMR].

Following the recommendations of Howe et.al (2021)[18] and Burgess and Labrecque (2018)[19], for the analyses exploring the effect of genetic liability to Income and EA on the risk of mental disorders, causal effects and 95% confidence intervals (CI) were expressed per one-SD increase in the odds of developing each mental disorder. For analyses exploring the effect of mental disorders on EA and income, we multiplied the causal estimates by the standard deviation (SD) of income (SD= 33,181£)[5] and EA (SD= 4.2 years of education), then multiplied by 0.693 (i.e., ln2) to express the effect of a doubling of genetic liability to each mental disorders on the genetic liability to income or EA.

Finally, we used the considered significance after multiple testing correction (FDR < 0.05) to compare each sensitivity analysis across the same SES or Mental disorder phenotype pairs. For the MVMR analyses we have considered as significant only results included in confident intervals after non-parametric 1000 iteration bootstrap permutation. All the analyses were performed with R version 4.1.2

**eTable 1. Univariable and Multivariable mendelian randomisation results of socioeconomic status phenotypes on mental disorders.**

**eTable 1A: The total and direct (not mediated by educational attainment) effect estimates of genetic liability to higher income on the risk of mental disorders**

| exposure | outcome | method | nsnp | or | se | 95% CI | pval | OR 1k bootstrap | 95%CI 1K bootstrap |
|---|---|---|---|---|---|---|---|---|---|
| Higher Income | Schizophrenia | IVW | 44 | 0.63 | 0.221 | 0.408, 0.973 | 0.0372 | | |
| **Higher Income (not mediated by EA)** | **Schizophrenia** | **IVW-MVMR** | **298** | **0.385** | **0.214** | **0.218, 0.679** | **6.38E-04** | **0.322** | **0.160, 0.624** |
| Higher Income | Bipolar disorder | IVW | 44 | 1.127 | 0.202 | 0.757, 1.677 | 0.555 | | |
| Higher Income (not mediated by EA) | Bipolar disorder | IVW-MVMR | 298 | 0.543 | 0.298 | 0.303, 0.975 | 0.042 | 0.592 | 0.253, 1.472 |
| **Higher Income** | **MDD** | **IVW** | **41** | **0.669** | **0.075** | **0.577, 0.775** | **9.82E-08** | | |
| **Higher Income (not mediated by EA)** | **MDD** | **IVW-MVMR** | **295** | **0.661** | **0.104** | **0.539, 0.812** | **9.96E-05** | **0.673** | **0.577, 0.775** |

**eTable 1B: The total and direct (not mediated by income) effect estimates of genetic liability to higher educational attainment on the risk of mental disorders**

| exposure | outcome | method | nsnp | or | se | 95%_CI | pval | OR 1k bootstrap | 95% CI 1K bootstrap |
|---|---|---|---|---|---|---|---|---|---|
| Higher Educational Attainment | Schizophrenia | IVW | 302 | 1.115 | 0.098 | 0.920, 1.352 | 0.263 | | |
| **Higher Educational Attainment (not mediated by income)** | **Schizophrenia** | **IVW-MVMR** | **298** | **2.095** | **0.289** | **1.374, 3.189** | **1.09E-03** | **2.513** | **1.485, 4.195** |

| | | | | | | | | | |
|---|---|---|---|---|---|---|---|---|---|
| **Higher Educational Attainment** | **Bipolar disorder** | **IVW** | **283** | **1.841** | **0.114** | **1.501, 2.257** | **4.81E-09** | | |
| **Higher Educational Attainment (not mediated by income)** | **Bipolar disorder** | **IVW-MVMR** | **298** | **2.694** | **0.104** | **1.747, 4.156** | **1.14E-05** | **2.262** | **1.008, 4.246** |
| **Higher Educational Attainment** | **MDD** | **IVW** | **279** | **0.783** | **0.038** | **0.726, 0.844** | **1.89E-10** | | |
| Higher Educational Attainment (not mediated by income) | MDD | IVW-MVMR | **295** | 1.027 | 0.077 | 0.881, 1.196 | 0.726 | 0.994 | 0.847, 1.180 |

#### eTable 2. Univariable Mendelian Randomisation Results of Educational attainment Versus Severe mental Disorders and sensitivity analyses

#### eTable 2A. Univariable Mendelian Randomisation Results of Educational attainment Versus Severe mental Disorders

| exposure | F stat | outcome | method | nsnp | b | se | pval | Low ci | Up ci | OR | Or low CI | Or up ci |
|---|---|---|---|---|---|---|---|---|---|---|---|---|
| Educational Attainment | 29.72-239.82 | Schizophrenia | MR Egger | 302 | 0.696829 | 0.387397 | 0.07306369 | -0.0624687 | 1.45612 | 2.0073764 | 0.9394424 | 4.28931 |
| | | | Weighted median | 302 | -0.02595 | 0.083958 | 0.75724799 | -0.1905088 | 0.13860 | 0.9743827 | 0.8265384 | 1.148672 |
| | | | Inverse variance weighted | 302 | 0.109735 | 0.098119 | 0.26340141 | -0.0825784 | 0.302049 | 1.1159828 | 0.9207392 | 1.352628 |
| | | | Simple mode | 302 | -0.00577 | 0.334652 | 0.98624344 | -0.6616933 | 0.650144 | 0.9942417 | 0.5159769 | 1.915816 |
| | | | Weighted mode | 302 | -0.03672 | 0.298831 | 0.9022831 | -0.6224302 | 0.548989 | 0.9639452 | 0.5366387 | 1.731501 |
| | | bipolar disorder | MR Egger | 283 | 0.346463 | 0.411465 | 0.400491152 | -0.4600087 | 1.152935 | 1.414057486 | 0.631278122 | 3.167476 |
| | | | **Weighted median** | **283** | **0.492999** | **0.114745** | **1.73538E-05** | **0.2680983** | **0.717899** | **1.637218122** | **1.307475764** | **2.050121** |
| | | | **Inverse variance weighted** | **283** | **0.610118** | **0.104228** | **4.80729E-09** | **0.40583179** | **0.814404** | **1.840648206** | **1.500550127** | **2.257829** |

| | | | | | | | | | | | | |
|---|---|---|---|---|---|---|---|---|---|---|---|---|
| | | | Simple mode | 283 | 0.225323 | 0.46803 | 0.6305849 | -0.6920156 | 1.142662 | 1.252727666 | 0.5005661 | 3.135104 |
| | | | Weighted mode | 283 | 0.196803 | 0.477648 | 0.680634222 | -0.7393872 | 1.132993 | 1.21750398 | 0.477406369 | 3.104935 |
| | | Major depression disorder | MR Egger | 279 | -0.11739 | 0.151237 | 0.43829301 | -0.4138148 | 0.179034 | 0.889238124 | 0.661123371 | 1.196062 |
| | | | **Weighted median** | **279** | **-0.2444** | **0.03821** | **1.60148E-10** | **-0.3193072** | **-0.1695** | **0.783170894** | **0.726652251** | **0.844086** |
| | | | **Inverse variance weighted** | **279** | **-0.24383** | **0.03828** | **1.89424E-10** | **-0.3188637** | **-0.1688** | **0.783617631** | **0.726974592** | **0.844674** |
| | | | Simple mode | 279 | -0.42569 | 0.12457 | 0.000727329 | -0.6698533 | -0.18153 | 0.653318992 | 0.511783625 | 0.833996 |
| | | | Weighted mode | 279 | -0.39116 | 0.122832 | 0.00161485 | -0.6319118 | -0.15041 | 0.676271189 | 0.531574549 | 0.860355 |

#### eTable 2B. Sensitivity analyses of Educational attainment Versus Severe mental Disorders and sensitivity analyses

| exposure | outcome | Het method | Q | Q df | Q pval | Egger intercept | Pleo se | Pleo pval | MR-PRESSO Outliers | MR-PRESSO Beta-corrected | MR-PRESSO Sd | MRPRESSO Pval |
|---|---|---|---|---|---|---|---|---|---|---|---|---|
| Educational Attainment | Schizophrenia | Inverse variance weighted | 1656.253 | 301 | 1.05E-185 | -0.0082675 | 0.005278343 | 0.118331 | 32 | 0.04645835 | 0.074802 | 0.535068 |
| | bipolar disorder | Inverse variance weighted | 674.4342 | 282 | 3.62E-34 | 0.003711469 | 0.00560291 | 0.508246 | **5** | **0.5352** | **0.0989** | **1.35E-07** |
| | Major depression disorder | Inverse variance weighted | 866.293 | 278 | 1.17E-61 | -0.00177775 | 0.002057027 | 0.388208 | **14** | **-0.2119** | **0.0311** | **6.68E-11** |

#### eTable 3. Univariable Mendelian Randomisation Results of income Versus Severe mental Disorders and sensitivity

#### eTable 3A. Univariable Mendelian Randomisation Results of income Versus Severe mental Disorders

| exposure | F stat | outcome | method | nsnp | beta | se | pval | Low ci | Up ci | OR | Or low ci | Or up ci |
|---|---|---|---|---|---|---|---|---|---|---|---|---|
| Average total household income before tax | 29.965-101.391 | Schizophrenia | MR Egger | 44 | 0.58728 | 1.013924 | 0.565538 | -1.40001 | 2.574570 | 1.7990883 | 0.246595 | 13.1256748 |
| | | | Weighted median | 44 | -0.29674 | 0.150362 | 0.048437 | -0.59145 | -0.002032 | 0.7432361 | 0.553524 | 0.9979697 |
| | | | Inverse variance weighted | 44 | -0.46078 | 0.221204 | 0.037248 | -0.89434 | -0.027215 | 0.6307943 | 0.408879 | 0.973152 |
| | | | Simple mode | 44 | -0.43435 | 0.328264 | 0.192771 | -1.07775 | 0.209047 | 0.6476858 | 0.340362 | 1.2325035 |
| | | | Weighted mode | 44 | -0.41574 | 0.281634 | 0.147184 | -0.96774 | 0.136262 | 0.6598511 | 0.379939 | 1.1459825 |
| Average total household income before tax | | bipolar disorder | MR Egger | 44 | 0.15338 | 0.872481 | 0.861298 | -1.55668 | 1.863442 | 1.16576792 | 0.210834 | 6.44589148 |
| | | | Weighted median | 44 | 0.129797 | 0.196442 | 0.50878 | -0.25523 | 0.514822 | 1.138596898 | 0.774739 | 1.673342112 |
| | | | Inverse variance weighted | 44 | 0.119472 | 0.202879 | 0.555938 | -0.27817 | 0.517114 | 1.12690213 | 0.757168 | 1.677180793 |
| | | | Simple mode | 44 | 0.192195 | 0.459462 | 0.677807 | -0.70835 | 1.092740 | 1.211906555 | 0.492456 | 2.982435151 |
| | | | Weighted mode | 44 | 0.173875 | 0.455829 | 0.704749 | -0.71955 | 1.067299 | 1.189907128 | 0.486972 | 2.907518464 |
| Average total household income before tax | | Major depression | MR Egger | 41 | 0.087667 | 0.368594 | 0.813249 | -0.63478 | 0.810111 | 1.091624463 | 0.530053 | 2.248158006 |
| | | | **Weighted median** | **41** | **-0.26452** | **0.070541** | **0.000177** | **-0.40278** | **-0.126259** | **0.767575009** | **0.66846** | **0.881386502** |
| | | | **Inverse variance weighted** | **41** | **-0.40165** | **0.075357** | **9.82E-08** | **-0.54935** | **-0.253955** | **0.669211852** | **0.577323** | **0.775726348** |

| | | | | | | | | | | | |
|---|---|---|---|---|---|---|---|---|---|---|---|
| | | | Simple mode | 41 | -0.4595 | 0.182685 | 0.016013 | -0.81757 | -0.101440 | 0.631597313 | 0.441505 | 0.903534727 |
| | | | Weighted mode | 41 | -0.0533 | 0.150301 | 0.724732 | -0.34789 | 0.241289871 | 0.948094744 | 0.706175 | 1.272889956 |

#### eTable 3B. Sensitivity analyses of income Versus Severe mental Disorders and sensitivity analyses

| exposure | outcome | Het method | Q | Q df | Q pval | egger intercept | Pleo se | Pleo pval | MR-PRESSO Outliers | MR-PRESSO Beta-corrected | MR-PRESSO Sd | MRPRESSO Pval |
|---|---|---|---|---|---|---|---|---|---|---|---|---|
| Average total household income before tax | Schizophrenia | Inverse variance weighted | 356.6899 | 43 | **5.07E-51** | -0.0207 | 0.019547 | 0.2956063 | **11** | **-0.4108** | **0.142** | **0.006913** |
| Average total household income before tax | bipolar disorder | Inverse variance weighted | 122.7679 | 43 | **1.33E-09** | -0.00068 | 0.016958 | 0.968293805 | 3 | -0.109 | 0.157 | 0.491 |
| Average total household income before tax | Major depression | Inverse variance weighted | 143.714 | 40 | **1.29E-13** | -0.00951 | 0.007016 | 0.183033518 | 5 | **-0.315** | **0.058** | **5.47E-06** |

#### eTable 4. Univariable Mendelian Randomisation Results and sensitivity analyses of genetic liability to severe mental disorders Versus Educational attainment

#### eTable 4A. Univariable Mendelian Randomisation Results of genetic liability to severe mental disorders Versus Educational attainment

| exposure | F stat | outcome | method | nsnp | beta | se | pval | low ci | up ci | or | or low ci | or up ci | months | low ci months | up ci months |
|---|---|---|---|---|---|---|---|---|---|---|---|---|---|---|---|
| Schizophrenia | 29.76 174.94 | Educational attainment | MR Egger | 145 | 0.038267316 | 0.028906 | 0.187657 | -0.01839 | 0.094922 | 1.039009 | 0.9817805 | 1.099573 | 1.3259625 | -0.6371271 | 3.289052 |
| | | | Weighted median | 145 | 0.000924741 | 0.005021 | 0.853884 | -0.00892 | 0.010766 | 1.000925 | 0.9911227 | 1.010825 | 0.0320422 | -0.308971 | 0.373056 |
| | | | Inverse variance weighted | 145 | 0.002087898 | 0.007425 | 0.778559 | -0.01247 | 0.016641 | 1.00209 | 0.9876122 | 1.01678 | 0.0723456 | -0.4319182 | 0.57661 |
| | | | Simple mode | 145 | -0.00215368 | 0.014211 | 0.879751 | -0.03001 | 0.025699 | 0.997849 | 0.9704392 | 1.026032 | -0.0746249 | -1.0397246 | 0.890475 |
| | | | Weighted mode | 145 | -0.00352479 | 0.010202 | 0.730233 | -0.02352 | 0.016472 | 0.996481 | 0.976753 | 1.016608 | -0.1221340 | -0.8150208 | 0.570753 |
| bipolar disorder | 29.75-56.56 | | MR Egger | 13 | 0.176621664 | 0.101688 | 0.110287 | -0.02269 | 0.375931 | 1.19318 | 0.977568219 | 1.456345955 | 6.1199406 | -0.7861115 | 13.02599 |
| | | | Weighted median | 13 | 0.018245019 | 0.009159 | **0.046379** | 0.000292 | 0.036198 | 1.018412 | 1.000292466 | 1.036860725 | 0.6321899 | 0.01013247 | 1.254247 |
| | | | Inverse variance weighted | 13 | 0.038493463 | 0.017144 | **0.024749** | 0.004891 | 0.072096 | 1.039244 | 1.004903054 | 1.074758357 | 1.3337984 | 0.16947569 | 2.498121 |
| | | | Simple mode | 13 | 0.020641715 | 0.012462 | 0.123544 | -0.00378 | 0.045067 | 1.020856 | 0.996223181 | 1.046098365 | 0.71523544 | -0.1311145 | 1.561585 |
| | | | Weighted mode | 13 | 0.020641715 | 0.012017 | 0.111527 | -0.00291 | 0.044195 | 1.020856 | 0.99709218 | 1.045186655 | 0.71523544 | -0.1009027 | 1.531374 |
| Major depression | 29.83-77.80 | | MR Egger | 44 | 0.093009723 | 0.188072 | 0.6235 | -0.27561 | 0.461631 | 1.097472 | 0.759107938 | 1.586659318 | 3.22278691 | -9.5499316 | 15.99551 |
| | | | **Weighted median** | **44** | **-0.04471068** | **0.019727** | **0.023424** | **-0.08338** | **-0.00605** | **0.956274** | **0.920005219** | **0.993972809** | **-1.54922512** | **-2.8889762** | **-0.20947** |
| | | | **Inverse variance weighted** | **44** | **-0.0807963** | **0.035011** | **0.021015** | **-0.14942** | **-0.01217** | **0.922382** | **0.861208583** | **0.987899744** | **-2.79959192** | **-5.1773527** | **-0.42183** |
| | | | Simple mode | 44 | -0.03859987 | 0.035554 | 0.283669 | -0.10828 | 0.031085 | 0.962136 | 0.897371862 | 1.031573394 | -1.33748544 | -3.7520732 | 1.077102 |
| | | | Weighted mode | 44 | -0.04463461 | 0.028262 | 0.121594 | -0.10003 | 0.010759 | 0.956347 | 0.904811754 | 1.010817236 | -1.546589166 | -3.4659828 | 0.372804 |

**eTable 4B. Sensitivity analyses of genetic liability to severe mental disorders Versus Educational attainment**

| exposure | outcome | het method | Q | Q df | Q pval | Egger intercept | Pleo se | Pleo pval | MR-PRESSO Outliers | MR-PRESSO Beta-corrected | MR-PRESSO Sd | MRPRESSO Pval |
|---|---|---|---|---|---|---|---|---|---|---|---|---|
| Schizophrenia | Educational attainment | Inverse variance weighted | 1338.781 | 144 | **1.07E-192** | -0.00246 | 0.001897 | 0.197446 | 26 | 0.010340794 | 0.004817632 | **0.03388456** |
| bipolar disorder | Educational attainment | Inverse variance weighted | 98.58339 | 12 | **1.05E-15** | -0.0131 | 0.009512 | 0.195985 | 4 | 0.011 | 0.006 | 0.1051 |
| Major depression | Educational attainment | Inverse variance weighted | 580.934 | 43 | **2.15E-95** | -0.00537 | 0.005713 | 0.352271 | **15** | **-0.055** | **-2.946** | **0.0064** |

#### eTable 5. Univariable Mendelian Randomisation Results and sensitivity analyses of genetic liability to severe mental disorders Versus household income

#### eTable 5A. Univariable Mendelian Randomisation Results of genetic liability to severe mental disorders Versus household income

| exposure | F stat | outcome | method | nsnp | beta | se | pval | low ci | up ci | or | or low ci | or up ci | months | low ci months | up ci months |
|---|---|---|---|---|---|---|---|---|---|---|---|---|---|---|---|
| Schizophrenia | 29.76- 174.94 | household income | MR Egger | 149 | -0.0122 | 0.024737 | 0.622595 | -0.06069 | 0.036284 | 0.987873 | 0.941119 | 1.036951 | -280.553 | -1395.45 | 834.3397 |
| | | | **Weighted median** | **149** | **-0.02522** | **0.006674** | **0.000158** | **-0.0383** | **-0.01214** | **0.975097** | **0.962424** | **0.987937** | **-579.883** | **-880.686** | **-279.08** |
| | | | **Inverse variance weighted** | **149** | **-0.03566** | **0.006632** | **7.55E-08** | **-0.04866** | **-0.02266** | **0.964968** | **0.952506** | **0.977592** | **-819.999** | **-1118.88** | **-521.123** |
| | | | Simple mode | 149 | -0.01316 | 0.018142 | 0.469454 | -0.04871 | 0.022401 | 0.986929 | 0.952453 | 1.022654 | -302.538 | -1120.17 | 515.094 |
| | | | Weighted mode | 149 | -0.02387 | 0.015461 | 0.124767 | -0.05417 | 0.006434 | 0.976414 | 0.947269 | 1.006455 | -548.85 | -1245.66 | 147.9571 |

| | | | | | | | | | | | | | | | |
|---|---|---|---|---|---|---|---|---|---|---|---|---|---|---|---|
| bipolar disorder | 29.75- 56.56 | | MR Egger | 13 | 0.131272 | 0.076593 | 0.114552 | -0.01885 | 0.281395 | 1.140278 | 0.981326 | 1.324977 | 3018.527 | -433.464 | 6470.519 |
| | | | Weighted median | 13 | 0.020942 | 0.013706 | 0.126523 | -0.00592 | 0.047805 | 1.021163 | 0.994096 | 1.048966 | 481.5426 | -136.159 | 1099.245 |
| | | | Inverse variance weighted | 13 | 0.015913 | 0.013129 | 0.225485 | -0.00982 | 0.041645 | 1.01604 | 0.990229 | 1.042525 | 365.9094 | -225.791 | 957.6095 |
| | | | Simple mode | 13 | 0.025016 | 0.026862 | 0.370064 | -0.02763 | 0.077665 | 1.025331 | 0.972745 | 1.08076 | 575.2256 | -635.406 | 1785.857 |
| | | | Weighted mode | 13 | 0.028137 | 0.024136 | 0.266346 | -0.01917 | 0.075444 | 1.028537 | 0.981014 | 1.078362 | 647.005 | -440.774 | 1734.784 |
| | | | MR Egger | 44 | -0.02692 | 0.183566 | 0.884093 | -0.38671 | 0.332866 | 0.973435 | 0.679285 | 1.394961 | -619.1 | -8892.27 | 7654.072 |
| Major depression | 29.83-77.80 | | **Weighted median** | **44** | **-0.1283** | **0.02625** | **1.02E-06** | **-0.17975** | **-0.07685** | **0.87959** | **0.83548** | **0.926028** | **-2950.18** | **-4133.23** | **-1767.13** |
| | | | **Inverse variance weighted** | **44** | **-0.11478** | **0.034471** | **0.00087** | **-0.18234** | **-0.04721** | **0.891566** | **0.83332** | **0.953884** | **-2639.19** | **-4192.75** | **-1085.63** |
| | | | Simple mode | 44 | -0.13193 | 0.06014 | 0.033703 | -0.24981 | -0.01406 | 0.876399 | 0.778951 | 0.986038 | -3033.74 | -5744.18 | -323.307 |
| | | | Weighted mode | 44 | -0.13514 | 0.054201 | 0.016584 | -0.24137 | -0.0289 | 0.873594 | 0.785548 | 0.971509 | -3107.45 | -5550.26 | -664.642 |

#### eTable 5B. Sensitivity analyses of genetic liability to severe mental disorders Versus household income

| exposure | outcome | het_method | Q | Q_df | Q_pval | egger_intercept | pleo_se | pleo_pval | **MR-PRESSO Outliers** | **MR-PRESSO Beta-corrected** | **MR-PRESSO Sd** | **MRPRESSO Pval** |
|---|---|---|---|---|---|---|---|---|---|---|---|---|
| Schizophrenia | household income | Inverse variance weighted | 268.7262 | 68 | **1.15E-25** | -0.0016 | 0.001641 | 0.326539 | **9** | **-0.0254** | **0.00546** | **7.2015e-06** |
| bipolar disorder | household income | Inverse variance weighted | 23.74166 | 12 | 0.02205 | -0.01095 | 0.007171 | 0.155125 | 1 | 0.0241 | 0.0106 | **0.0435** |

| | | | | | | | | | | | | |
|---|---|---|---|---|---|---|---|---|---|---|---|---|
| Major depression | household income | Inverse variance weighted | 233.2531 | 43 | **5.72E-28** | -0.00272 | 0.005585 | 0.628503 | **6** | **-0.0977** | **0.0254** | **0.00046** |

#### eTable 6. Multivariable Mendelian Randomisation Results of Socioeconomic status and Intelligence versus severe mental disorders

| exposure | Conditional F-statistics | outcome | nsnp | OR | beta | Beta low ci | Beta up ci | se | pval | Modified Q-Statistic | Q pval | Robust Beta 1k bootstrap | Low 95% CI 1k bootstrap | Up 95% CI 1k bootstrap |
|---|---|---|---|---|---|---|---|---|---|---|---|---|---|---|
| **Intelligence** | **13.91762** | **Schizophrenia** | **88** | **0.66729** | **-0.40453** | **-0.68984** | **-0.11923** | **0.145564** | **5.45E-03** | 1520.264 on 325 DF | **1.82e-153** | **-0.51422** | **-0.789** | **-0.246** |
| **Educational Attainment** | **8.009355** | **Schizophrenia** | **268** | **3.19179** | **1.160582** | **0.70675** | **1.614414** | **0.231547** | **5.38E-07** | | | **1.23484** | **0.768** | **1.815** |
| **household income** | **5.133689** | **Schizophrenia** | **21** | **0.32877** | **-1.1124** | **-1.65816** | **-0.56663** | **0.27845** | **6.47E-05** | | | **-1.06974** | **-1.713** | **-0.476** |
| **Intelligence** | **13.91762** | **bipolar disorder** | **88** | **0.673189** | **-0.39573** | **-0.70435** | **-0.08711** | **0.157461** | **0.011964** | 777.7144 on 325 DF | **4.165e-39** | **-0.43973** | **-0.902** | **-0.086** |
| **Educational Attainment** | **8.009355** | **bipolar disorder** | **268** | **3.607148** | **1.282917** | **0.791763** | **1.774072** | **0.250589** | **3.06E-07** | | | **1.158732** | **0.436** | **1.806** |
| household income | **5.133689** | bipolar disorder | 21 | 0.563794 | -0.57307 | -1.16372 | 0.017591 | 0.301356 | 0.05722 | | | -0.47143 | -1.273 | 0.633 |
| **Intelligence** | **13.91762** | **Major depression** | **88** | **1.25118** | **0.224087** | **0.118784** | **0.329391** | **0.053726** | **3.03E-05** | 844.850 on 323 DF | **2.546e-48** | **0.160596** | **0.025** | **0.282** |
| Educational Attainment | **8.009355** | Major depression | 266 | 0.888986 | -0.11767 | -0.28549 | 0.050141 | 0.085619 | 0.169325 | | | -0.09043 | -0.296 | 0.086 |
| **household income** | **5.133689** | **Major depression** | **21** | **0.639582** | **-0.44694** | **-0.64866** | **-0.24522** | **0.102919** | **1.41E-05** | | | **-0.44459** | **-0.682** | **-0.208** |

*Bold (MVMR with 95% confidence intervals included after non-parametric 1000 iteration bootstrap permutations)*
